# Surges in COVID-19 are led by lax government interventions in initial outbreaks

**DOI:** 10.1101/2020.07.17.20156604

**Authors:** Hsiang-Yu Yuan, Lindsey Wu, Dong-Ping Wang

**Affiliations:** Department of Biomedical Sciences, Jockey Club College of Veterinary Medicine and Life Sciences, City University of Hong Kong, Hong Kong; Department of Infection Biology, Faculty of Infectious and Tropical Diseases, London School of Hygiene & Tropical Medicine, United Kingdom; School of Marine and Atmospheric Sciences, Stony Brook University, Stony Brook, New York, USA

## Abstract

Sharp increases in COVID-19 cases occurred after reopening in the United States. We show that the post-intervention effective reproduction number is a strong predictor of the surge in late June. Lax interventions in the early stages coupled with elevated virus spread are primarily responsible for surges in most affected states.

## Main text

In the United States, there were over 3 million confirmed cases of COVID-19 by July 6. The initial major outbreak, centered in New York, had begun in mid-March, and reached an apex by early April. As the states opened up, the daily counts have spiked since late June. Several big states, which had avoided large initial outbreaks, e.g., California, Florida, and Texas, are the hardest hit. These states may have ended lockdowns too soon and/or relaxed restrictions too aggressively. We address the specific question whether lax control measures during initial outbreaks may have played a key role in the current surge.

We obtained the daily new cases from “The COVID Tracking Project”, https://covidtracking.com/, for the fourteen states of among the highest total cases in the United States. The study period is from the date of the first reported cases in each state up to July 6. The total cases *C(t)* tend to increase exponentially with a growth rate such that the number of daily new cases is Δ*C* = *λC*. The growth rate is a measure of the percentage increase of daily new cases, commonly used to describe the virus spread *(1)*. To capture the current outbursts, we calculated the two-week averaged growth rates for each state using data from the last 14 days of records (Fig. 1A). Arizona, Florida, and Texas stood out with the highest growth rates > 0.04 days^-1^. California, North Carolina, Georgia, and Louisiana also had fast growth rates. In contrast, New York, New Jersey, and Massachusetts, had the lowest growth rates < 0.002 days^-1^.

**Figure 1.**
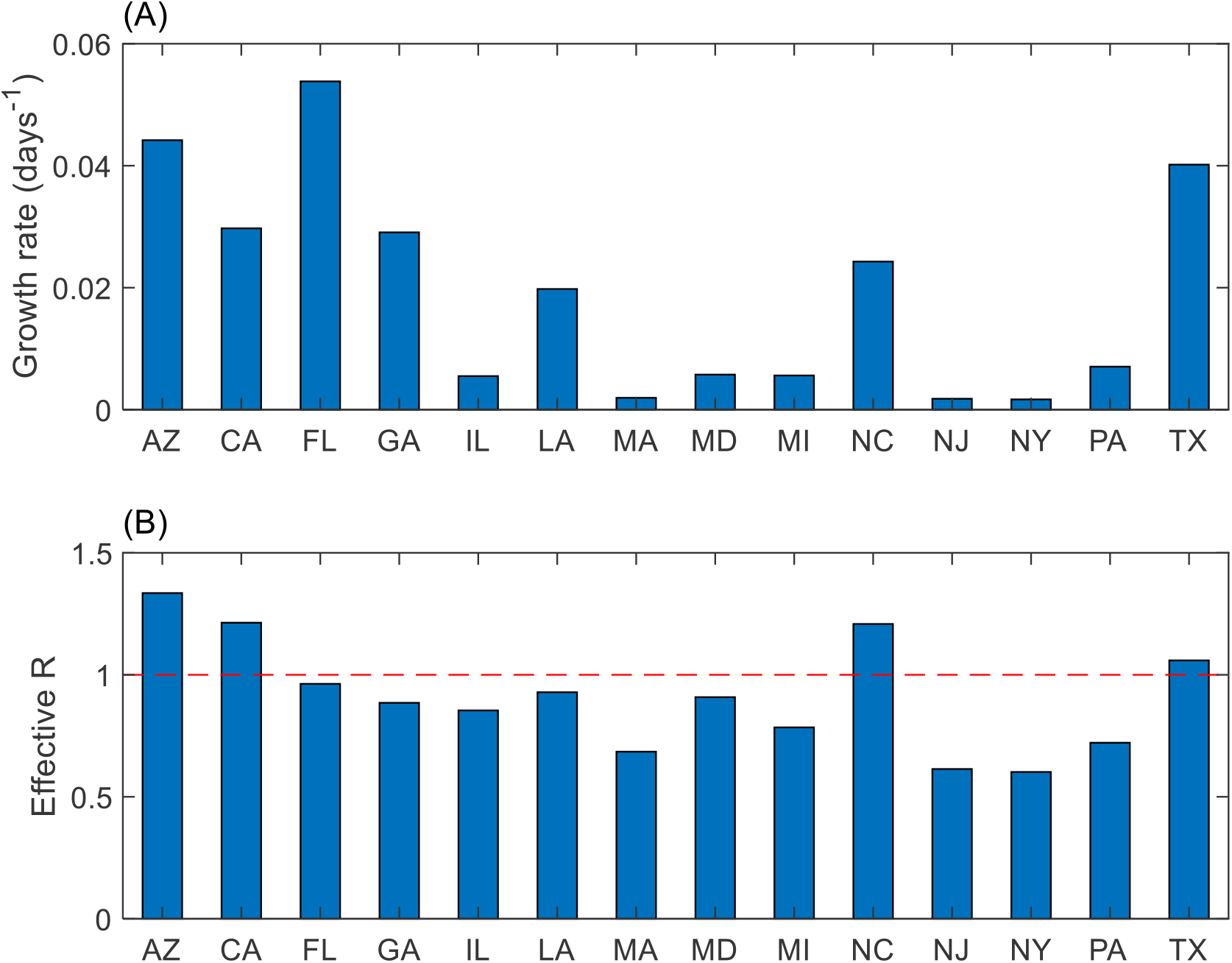
The exponential growth rates in late June calculated from the observations (A); and the effective reproduction number estimated from a model-filter system using the first 91 days of observations (B). The horizontal red dashed line marks the critical value (*R*_*e*_ = 1).

To explain the pattern of observed growth rates, a susceptible-infectious-recovered (SIR) model is used to estimate the effective reproduction number *R*_*e*_ in early June prior to the current surge. To account for government interventions, the model includes a time-dependent transmission rate such that after full interventions the effective reproduction number is *R*_*e*_ = *α R*_*o*_, where *R*_*o*_ is the basic reproduction number, and *α* <1, a measure of the effects of interventions. The parameter estimation is based on an ensemble Kalman filter (EnKF) *(2,3)* The model-filter system is applied for each state to obtain *R*_*e*_, using the first 91 days of the record (Fig. 1B). The posterior estimates of total cases are remarkable that the errors in most states were within 7% of the observed.

To successfully contain an epidemic, the effective reproduction number needs to stay beneath *R*_*e*_ = 1 *(4)*. Yet, *R*_*e*_ in early June were already above 1 for Arizona, California, North Carolina, and Texas, and were near 1 for Florida, Georgia, and Louisiana. All of these states experienced surges in late June. In contrast, the Northeast had the lowest *R*_*e*_ < 0.7, and the corresponding lowest growth rates. This result supports the hypothesis that the surges in late June were led by less effective early interventions evidenced by *R*_*e*_ above or near 1 in early June.

In the United States, each state has adopted a different policy towards lockdowns and reopening. In many states, the surges have often been attributed to early reopening. Arizona, Florida, Georgia, Louisiana, North Carolina, and Texas have all opened up (long) before the late May. The *R*_*e*_ above or near 1 in early June could result from premature relaxation of the lockdowns. Most of these states moreover have experienced 2-3 folds increases in growth rates in the month of June alone; the crowded bars and beaches may have been a key factor. The current crisis is thus a result of preexisting conditions due to relaxed initial interventions intensified by elevated virus spread rates since June.

California presents a seemingly contradictory case. The state didn’t open up till late May; yet, *R*_*e*_ was already above 1 in early June. After the reopening, the growth rate in California had only increased slightly. The surge thus is primarily associated with a persistent upward trend. At a growth rate of *λ* = 0.025 days^-1^ in early June (Fig. 1A), the daily counts would double in about a month. Indeed, the weekly averages had increased from about 2,600 in late May to an alarming 5,700 in late June. The current spike reflects a failure of initial interventions in fully containing the outbreak. North Carolina has a similar situation of an upward trend and no increased growth rate. Illinois and Maryland where *R*_*e*_ in early June were close to 1, offer another interesting case (Fig. 1B). The growth rates decreased considerably after the reopening, and both states have (so far) avoided a surge.

The post-intervention effective reproduction number provides a general measure of intervention outcomes, and is a robust indicator of the current surges. While the early reopening intensified the current crisis, it may not be the critical driver. To prevent resurgence, it is important to understand why in certain states, lockdowns did not lead to the anticipated effects. We suggest that behavioral factors related to compliance with public health measures, such as social distancing and face coverings, must be carefully considered *(5,6)*.

## Data Availability

Data are publicly available.

https://covidtracking.com/

## Acknowledgment

This study was supported by grants from the City University of Hong Kong (#7200573 and #9610416). We thank the support from the Centre for Applied One Health Research and Policy Advice, and Jockey Club College of Veterinary Medicine and Life Sciences, City University of Hong Kong.

## Notes

### Competing Interest Statement

The authors have declared no competing interest.

### Author Declarations

No IRB are needed.

